# Real-time Emotion Detection by Quantitative Facial Motion Analysis

**DOI:** 10.1101/2022.10.28.22276059

**Authors:** Jordan R. Saadon, Fan Yang, Ryan Burgert, Selma Mohammad, Theresa Gammel, Michael Sepe, Miriam Rafailovich, Charles B. Mikell, Pawel Polak, Sima Mofakham

## Abstract

**Background:** Research into mood and emotion has often depended on slow and subjective self-report, highlighting a need for rapid, accurate, and objective assessment tools.

**Methods:** To address this gap, we developed a method using digital image speckle correlation (DISC), which tracks subtle changes in facial expressions invisible to the naked eye, to assess emotions in real-time. We presented ten participants with visual stimuli triggering neutral, happy, and sad emotions and quantified their associated facial responses via detailed DISC analysis.

**Results:** We identified key alterations in facial expression (facial maps) that reliably signal changes in mood state across all individuals based on these data. Furthermore, principal component analysis of these facial maps identified regions associated with happy and sad emotions. Compared with commercial deep learning solutions that use individual images to detect facial expressions and classify emotions, such as Amazon Rekognition, our DISC-based classifiers utilize frame-to-frame changes. Our data show that DISC-based classifiers deliver substantially better predictions, and they are inherently free of racial or gender bias.

**Limitations:** Our sample size was limited, and participants were aware their faces were recorded on video. Despite this, our results remained consistent across individuals.

**Conclusions:** We demonstrate that DISC-based facial analysis can be used to reliably identify an individual’s emotion and may provide a robust and economic modality for real-time, noninvasive clinical monitoring in the future.

**Highlights:**

- We detected the emotional state of participants in the absence of visible changes in facial expressions
- We identified distinct facial movement maps for happy and sad emotions
- We developed a highly sensitive, noninvasive, noncontact emotional state assessment tool

## Introduction

The ability to accurately assess the internal states of the human mind remains one of the grand challenges of modern neuroscience. In recent years, considerable research has been devoted to discovering new methods for identifying moods and emotions. There are a number of tools used to assess mood psychometrically in the field of mood disorders research. Some are inventories, such as the Patient Health Questionairre-9 (PHQ-9) [1], the Remission Evaluation and Mood Inventory Tool (REMIT) [2], and the Ecological Momentary Assessment (EMA) [3], while others are symptom scales, including the Hamilton Rating Scale for Depression (HAM-D) [4], the Montgomery-Asburg Depression Rating Scale (MADRS) [5], and the Young Mania Rating Scale (YMRS) [6]. These tools are helpful in evaluating mood disorders in individuals with depressive or manic symptoms, but their validity and reliability are potentially compromised by methods of self-report or observer assessment [7–10]. Moreover, research into neurobiological mechanisms of mood requires temporal precision that survey-based instruments lack. The same applies to emotions. The subjectivity of existing scales has thus created a demand for objective measures of mood and emotional state.

Previous attempts at objective affective assessment have employed biosignal detection as a means of differentiating emotions. Some measures have targeted physiological markers of the stress response and emotional distress such as changes in facial skin temperature or color [11– 13]. Other studies have investigated existing diagnostic tools, such as electrocardiography [14], electroencephalography [15], and electromyography (EMG) [16,17]. Recently, the proliferation of wearable biosensor technology such as fitness trackers and smartwatches has yielded yet another potential tool for ambulatory mood assessment [18,19]. Analysis of the vast quantities of body-sensing data provided by such devices may be helpful in understanding the mechanistic foundations of mood and emotion. However, the validity and reliability of these measures compared to more traditional questionnaire-based methods have yet to be ascertained.

The face offers another promising avenue for real-time emotional assessment. Well over a century ago, William James observed that the affective state is usually reflected in facial movement (“… [the] neck is bent, the head hangs (‘bowed down’ with grief), the relaxation of the cheek- and jaw-muscles makes the face look long and narrow, the jaw may even hang open and the eyes appear large”) [20]. Since then, methods for correlating facial movement to the underlying emotional state were developed [21–23]. Popularized by Ekman and Friesen, the Facial Action Coding System (FACS) codifies facial movements based on the action units (muscles or groups of muscles) that create them. This system functions on the premise that the representation of emotions through facial movement are conserved across cultures and peoples [24]. Other studies have used facial EMG to detect purposeful changes in facial expression [25], as well as involuntary movements in response to affective touch [26]. Thus, quantitative analysis of facial movements may provide an intriguing modality for studying emotion and mood states.

A major drawback of the aforementioned methods is that they rely on detection of overt facial movements, however, individuals may attempt to conceal their emotions under certain circumstances. Even so, Ekman has argued that people unconsciously reveal their emotion through *microexpressions*, facial movements so brief that they are unrecognizable in real-time but can be decoded during close examination of videos [27,28]. Several methods have been developed to measure dynamic facial characteristics based on video recording of participants, including the central difference method [29]. The central difference method is an analytical approximation of a derivative, which in this case, Shreve and colleagues used to track the rate of change of facial movement as individuals displayed various emotions. This technique proved capable of detecting microexpressions, thus highlighting the potential utility of facial expression-based quantitative emotion detection methods. Whereas the authors of this prior study attempted to detect the occurrence of facial expressions [29], we sought to investigate whether we could predict an individual’s underlying affective state by simply looking at the face. The ability to ascertain an individual’s emotion or mood through facial analysis not only offers an objective tool for use in research settings, but also highlights the utility of such a tool in the clinical evaluation of affective disorders and monitoring treatment response.

Digital image speckle correlation (DISC) is a technique originally proposed for use in stress analysis of solid engineering materials. DISC tracks the geometric features of an object’s surface as it undergoes deformation [30]. More recently, researchers have discovered applications of this method to the fields of dermatology and reconstructive surgery [31–33]. By tracking the displacement of skin pores, DISC objectively quantifies facial movements in real-time [31]. Applying DISC to the human face has proved superior at facial recognition when compared with the traditional combined principal component analysis (PCA) and linear discriminant analysis method [32]. Other applications include studying changes in facial mechanical properties with aging [33] and after botulinum toxin treatment [34,35], as well as assessing facial nerve deficits in patients with vestibular schwannomas [36].

In the current study, we use DISC as a novel, real-time emotional assessment tool to track extremely subtle facial movements in order to reliably differentiate happy and sad emotions in healthy individuals. Unlike Ekman’s microexpressions, the minute changes in facial movement we observed are not discernable to the naked eye until after DISC analysis. Our results highlight the intriguing utility of this technique in emotion and mood detection as well as in more broad clinical monitoring settings.

## Methods

### Ethics Statement

This study was conducted under the supervision of the Stony Brook University Committee on Research in Human Subjects (IRB2019-0199). All volunteers gave their written informed consent before participation in the study.

### Participants and Videotaping

This pilot study included ten healthy volunteers (seven males and three females) aged 23-56 years (mean age: 31). Participants were initially instructed to fill out a baseline self-assessment manikin (SAM) form with respect to their current mood state. The SAM is a pictorial mood reporting method often used in conjunction with the international affective picture system (IAPS), the set of images used to elicit happy and sad moods in this study [37,38].

Study participants were asked to rest their chins on an apparatus consisting of a chinrest connected to a specially designed platform with a camera mount to keep the camera at a fixed distance from the face (**Supplementary Figure 1**). In this position, they were videotaped using a Canon EOS 60D camera while viewing an automated slideshow of images from the international affective picture system (IAPS). This set of images is designed to elicit emotional reactions in the viewer [37,38]. The slideshow consisted of ten consecutive images intended to elicit pleasant or happy emotions followed by ten consecutive images meant to evoke sad emotions; each image was shown for ten seconds. These images were selected at the discretion of the research team. A blank (white) screen was shown for ten seconds at the beginning, as well as between the two sets of images to provide the baseline facial expression. After image viewing, participants were instructed to fill out two additional SAMs, each with respect to their mood state while viewing each set of images.

### DISC analysis of facial movement

DISC was originally intended for stress analysis of various solid engineering materials [30]. By tracking geometric features of a specimen surface before and after deformation, DISC derives the corresponding displacements of the points on the surface. To track a point (pixel) with coordinates of (*x, y*) on the nondeformed image (**Supplementary Figure 2**), a neighborhood *N*_(*x, y*)_ of the pixel is defined that consists of a number of pixels, in which (*x, y*) are the coordinates of the center of that neighborhood. Then, this neighborhood is compared with an equal-sized one on the deformed image. Given the coordinates (*x*\*, *y*\*) of the center point of a neighborhood *N*_(*x*\*, *y*\*)_ on the deformed image, the similarity (S) of these two subsets can be evaluated using the cross-correlation function:

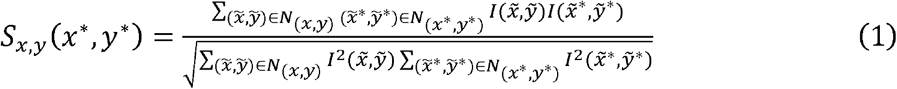

Where 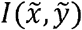 and 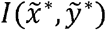 represent the gray-scale intensities (from 0 [black] to 255 [white]) of the corresponding pixels, and the summations are across the corresponding neighborhoods. DISC uses two frames as inputs and for every pixel with coordinates (*x, y*) in the first frame, it finds a pixel with coordinates (*x*\*, *y*\*) in the second frame with the highest similarity *S*_*x,y*_ (*x*\*, *y*\*). Therefore, DISC provides a displacement vector (*x*\*, *y*\*) - (*x, y*) for every pixel (*x, y*), yielding a vector field of displacement vectors for the whole image. This vector field characterizes movements on the specimen surface as defined by Peters and Ranson, and the length of the corresponding vectors corresponds to the intensity on the heatmaps used in our analysis [30].

By letting *u* and *v* correspond to the vertical and horizontal components of displacement, respectively:

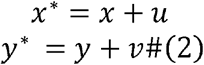

The displacement vector 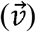 of the point (x, y) can be expressed as:

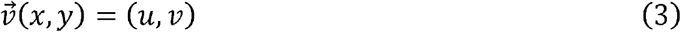

DISC analysis relies on tracking and characterizing “speckles” on the specimen surface [30]. As interpreted in Equation (1), each individual neighborhood within the image provides a distinct intensity profile. When applied to the human face, DISC utilizes skin pores that create ideal natural speckles to derive reliable displacement vectors of the face between two frames of a video [31,32]. Videos were split into individual frames. We categorized the frames based on the emotional valence (neutral, happy, or sad) of the image that the participant was viewing at that moment. One frame per second was used for analysis. Pairs of frames were analyzed via DISC^1^, with the first frame of both happy and sad image-viewing portions of the slideshow serving as the baseline to which all other happy and sad frames were compared. Using these frames of maximum proximity as a baseline minimized image misalignment, as participants may move their heads subtly throughout the slideshow. Resultant files containing displacement vectors for each point were then generated. Each point was located at the center of an 85 × 85-pixel subset with 20 pixels separating two given points.

### Heatmaps

Heatmaps in **Figures 1** and **2** demonstrate the varying magnitude of pixel displacement within each participant’s face. Heatmaps for both happy and sad emotions were generated from the average displacement throughout the viewing period for both happy and sad images. We then averaged these heatmaps from all participants to build composite heatmaps for happy and sad emotions and assess for spatial trends in facial expression changes (**Fig 3**). We analyzed the same number of pixels across individuals for comparison purposes.

**Fig 1^3^:** Heatmaps derived from the results of DISC analysis of representative frames for each emotion in a single participant. The top three panels are the original images, whereas the bottom three are the same images with superimposed heatmaps showing magnitude of movement from the baseline (neutral) frames. Units are in pixels.

**Fig 2^4^:** Heatmaps showing magnitude of facial movement in response to happy and sad images for each participant. Heatmaps were generated from the averaged DISC-calculated displacement across all happy and sad frames for that individual. Numbers represent each participant in the study. Participant 1 declined to have their face included in the publication of this data.

**Fig 3:**
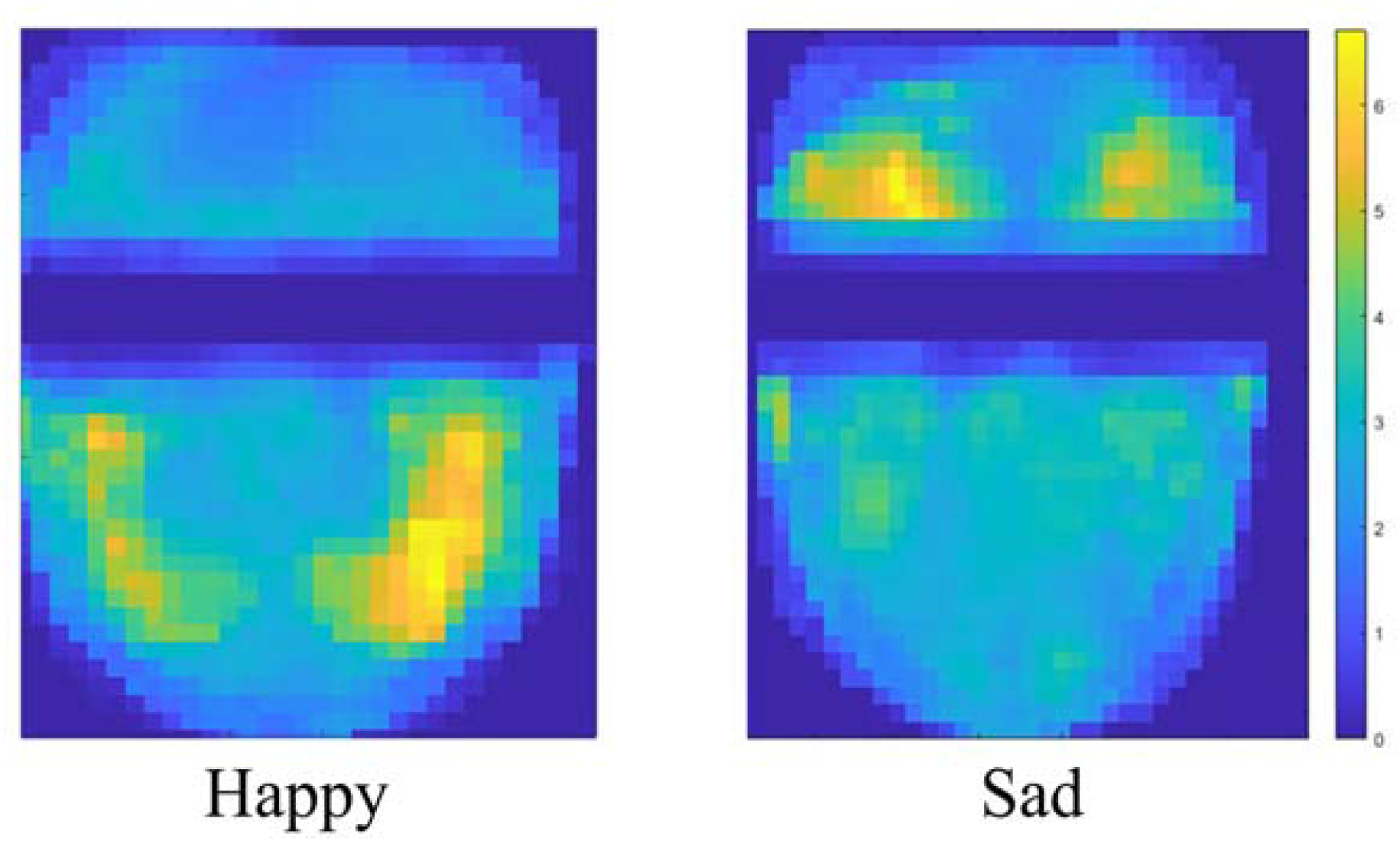
Average magnitude of facial movement in response to happy and sad images across all participants. Units are in pixels.

### Similarity Matrices

Frames from each participant’s video were individually compared, one against another, both within an emotion category and between emotions (e.g. happy-to-happy, sad-to-sad, and happy-to-sad), using the similarity values obtained from Equation (1). This was done to compare the frames of each video throughout the happy and sad image-viewing portions. Similarity values can range from zero (absence of any similarity) to one (identical frames). We then organized the calculated similarities into a matrix to visually demonstrate how similar each frame was to every other frame of a given participant. This was performed for all ten participants. To compare the average similarity among Happy-Happy and Sad-Sad quadrants to the Happy-Sad/Sad-Happy quadrants of each similarity matrix, we performed two-tailed t-tests using the SciPy library in Python.

### Principal Component Analysis

Principal component analysis (PCA) is a common method of converting high-dimensional data into lower-dimensional data. It is used to find a small subset of components that captures as much of the variance of the original data as possible [39]. In other words, it approximates the original information but in a compressed form. The principal components are defined as the eigenvectors of the covariance matrix of the original dataset, which in the context of this study, refers to a 500-dimensional vector for each frame of a video. For our analysis, we chose the two eigenvectors with the highest eigenvalues, as they are the two most informative components of the original, multidimensional vectors.

### Machine Learning-Based Mood Classifiers

Facial analysis is an intensive area of research. There exist numerous algorithms that detect, analyze, and read emotions from human faces. Big tech companies such as Amazon, Google, and Facebook develop many of these algorithms in-house and use them in their products. Chouinard and colleagues [40], evaluated several techniques in facial analysis primarily focusing on facial recognition and emotion detection. They concluded that Amazon Rekognition^2^ has the best performance for both face recognition and facial expression recognition. Amazon Rekognition is a commercial software that utilizes proprietary deep learning technology developed by Amazon Web Services (AWS). It is a static method, in other words, it takes an individual image as an input, and performs facial analysis—including the detection of eight different emotions (happy, surprised, confused, calm, angry, fear, disgusted, and sad).

Compared with static methods that use a single image, DISC employs a different set of predictors for categorizing emotion. It is a dynamic method that tracks subtle changes in facial movement in real-time between two frames of a video. In the final part of the next section, we demonstrate the out-of-sample performance of three different classifiers constructed with machine learning models that use DISC displacement data as features. We then compare these models to Amazon Rekognition.

The three classifiers are (i) a Multiclass Sparse Logistic Regression (MSLR); (ii) a Multi-Layer Perceptron (MLP); and (iii) a 3D-Convolutional Neural Network (CNN). MSLR is an extension of a well-known logistic regression method that allows for more than two categories of the predicted variable (see, e.g., Kim et al. 2006). It embeds feature selection into the classification framework using the ℓ_1_-norm regularization, and is attractive in many applications involving high-dimensional data. MLP is a class of feed-forward artificial neural networks from Pedregosa and colleagues (2011). It is a more flexible model than MSLR because it can capture nonlinear relationships between predictors. 3D-CNNs are a type of deep convolutional neural network that extracts features by performing 3D convolutions [41,42]. It captures the spatial information encoded in neighboring pixels in one heatmap, as well as the temporal information from the multiple adjacent frame heatmaps of a given participant.

The architecture of our 3D-CNN classifier is summarized in **Table S1**. It consists of two 3D convolution layers with a leaky ReLU activation function, max pooling in the convolution, and a batch normalization after each convolution for better numerical properties during the training of the network. Then, the network was flattened using global average pooling. It has two additional dense layers that shape the classifier into three final states that give the probabilities of each state. These probabilities were compared using a binary cross entropy function against the true labels (“Happy,” “Sad,” and “Neutral”). We experimented with different network architectures and shapes of the layers in the network. The out-of-sample results were robust against these modifications. Importantly, the confusion matrices and the classification errors were very similar for all the networks that we used.

## Results

### Facial Heatmaps

To visualize spatiotemporal facial expression changes associated with happy and sad emotions, we generated heatmaps demonstrating magnitude and direction of movement via DISC analysis of participant videos. The three images in the top panel of **Figure 1**, taken when the participant was exposed to neutral, happy, and sad-triggering stimuli, are virtually indistinguishable to the naked eye, making it difficult to discern what they are looking at. However, the differences among these affective states become more evident following DISC processing, where facial movement in response to happy and sad images localizes to distinct areas on the face, forming happy and sad facial maps. Importantly, these facial maps are conserved across participants. Examining the average heatmaps of all participants demonstrates a pattern whereby movement in response to happy images is concentrated in the lower face, around the angles of the mouth, and movement in response to sad images is concentrated in the brow area (**Fig 2**).

We next averaged facial movement across all participants to see if the patterns of happy and sad emotions observed earlier persisted (**Fig 3**). The composite heatmaps of average facial movement from all participants further demonstrated the differential localization of movement to the corners of the mouth during happy image-viewing and to the brow during sad image-viewing, consistent with our observations in **Figures 2** and **3**. Taken together, these results suggest that happy and sad images evoked subtle, yet spatially distinct changes in facial expression that are reliably detectable via DISC analysis and invariant of participants’ gender and age.

In order to assess whether a given participants responses to one image were consistent throughout the viewing period, we constructed similarity matrices. These similarity matrices compare the frames of each video throughout the happy and sad image-viewing portions to all other frames (**Fig 4**). Matrices were organized into four quadrants based on the category of the two frames being compared. Among all participants, the average similarity in both the Happy-Happy and Sad-Sad quadrants was significantly higher than that of the Happy-Sad and Sad-Happy quadrants (two-tailed t-test, p < 0.001). Therefore, these similarity matrices assert not only that the facial responses to happy and sad images are distinct, but that they are conserved across images.

**Fig 4:**
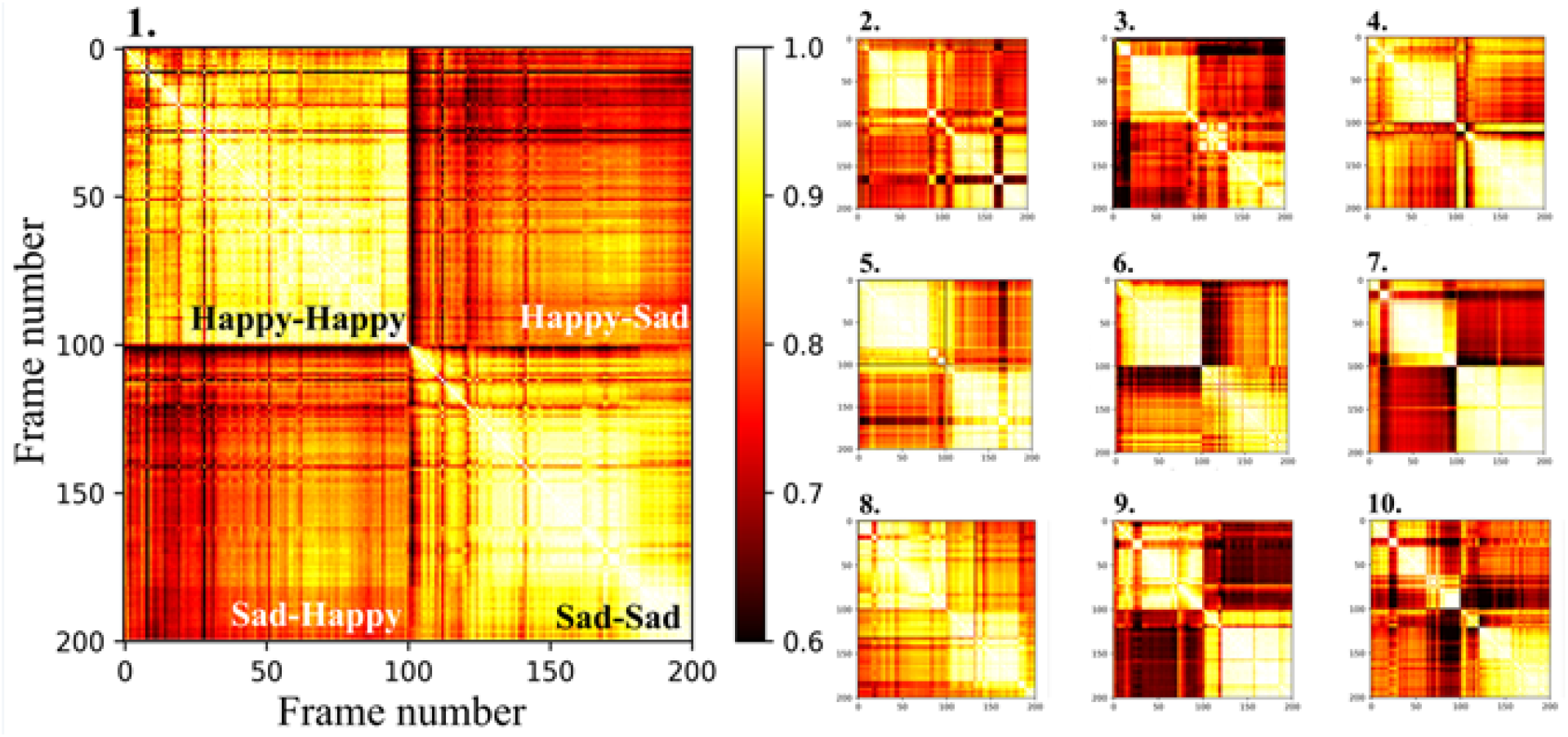
Similarity matrices of DISC results from each frame of each participant’s video. Matrices are numbered 1-10, corresponding to each participant’s ID. The matrix for Participant 1 is enlarged to show matrix organization. A value of 1.0 signifies 100% similarity between two frames and a value of 0 signifies the absence of any similarity.

### Spatial and Temporal Exploratory DISC Data Analysis

The use of PCA for facial recognition and analysis was first developed by Turk and Pentland [43]. Our files are large and high-dimensional, with each frame containing vectors with more than 500-dimensions. We applied PCA to the DISC-processed displacement data files to understand if a lower dimensional representation can still capture the salient mood information from our participants’ faces.

Dimensionality reduction via PCA is displayed in 2-D with every displacement vector represented as a pair of two numbers, corresponding to a point on the scatter plot shown in **Figure 5**. Although each frame contains greater than 500 displacement vectors associated with it, reducing this data to two values allows us to visually interpret our data while still representing over 50% of the total variance. Neutral frames cluster around the origin as they elicit almost no facial movement. This clustering also suggests that neutral frames are relatively similar within and across individuals. With respect to the non-neutral frames, a general trend exists whereby happy frames are more concentrated in the lower half of the plot and sad frames in the upper half. However, there is also a slight intermixing of happy and sad frames in the 2-D PCA. This observation demonstrates that although a gross trend separates these two emotions, a single frame’s first two principal components may not be sufficient to predict the participant’s affective state at that specific moment.

**Fig 5:**
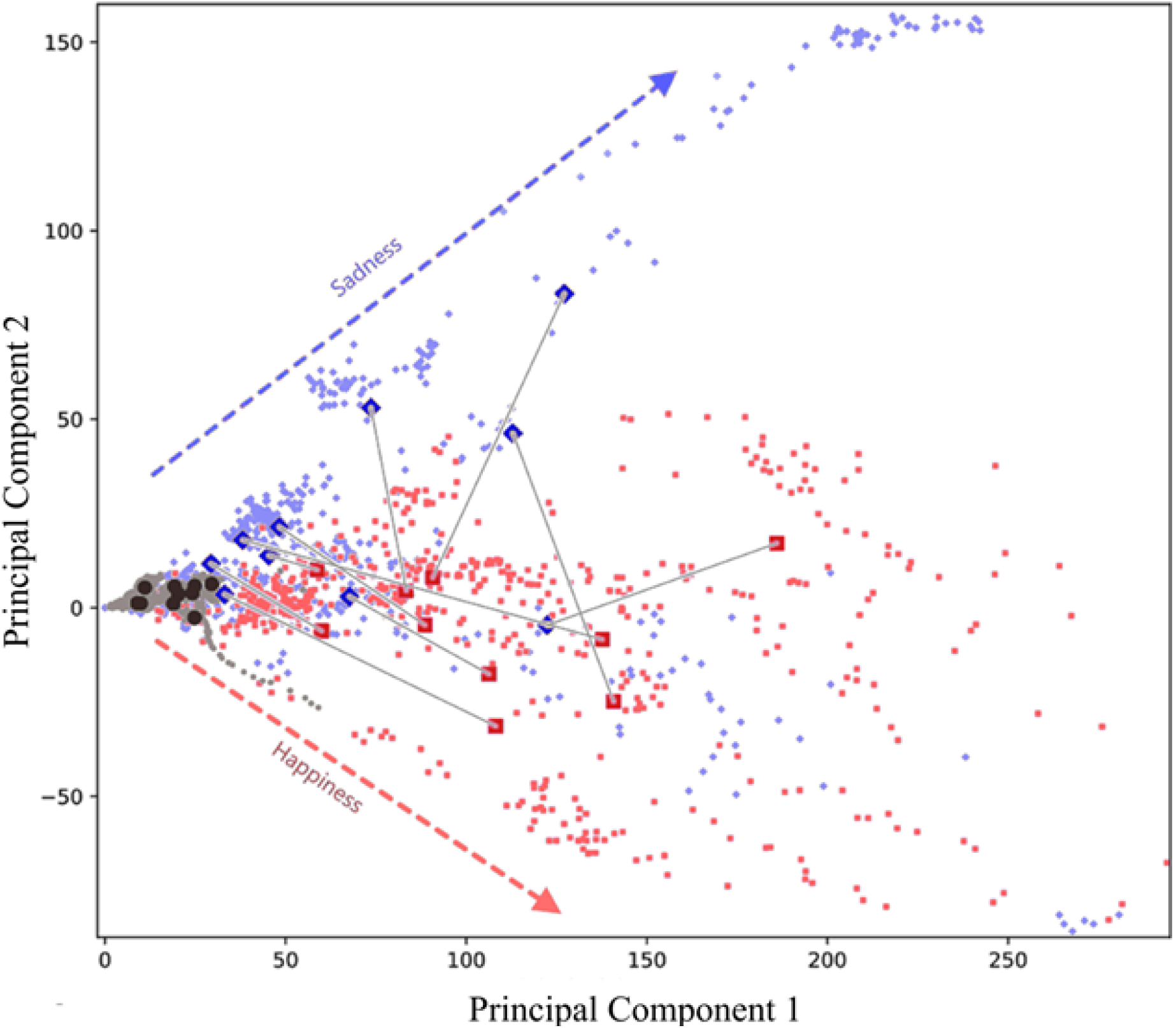
Plot of first and second principal components of DISC-processed displacement across all participants. Red squares represent frames from the happy image-viewing period, whereas blue diamonds represent frames from the sad image-viewing period. Black circles signify neutral frames. Large shapes indicate the averages for each participant. Gray lines serve to connect the averages for happy and sad for individual participants.

We reduced our data into two dimensions via PCA to not only compare our displacement data across participants, but also to determine whether the information captured by the first two principal components allows us to distinguish between happy and sad emotions. When considering the average principal component for each participant, two trends emerge. The first is that the happy frame average for all but one participant exhibited a larger first principal component than that of the sad frame average (**Fig 5**; rightward on the x-axis). Secondly, the sad frame average for all but one participant displays a higher second principal component than that of the average happy frame (upward on they-axis). Taken together, these trends suggest that PCA could distinguish the average DISC-detected facial movement in response to happy and sad images for eight of our ten participants.

Finally, because participants were successively shown a set of ten happy images followed by a set of ten sad images, we investigated the temporal features of facial movement during happy and sad emotions over the course of image presentation. The average of the magnitude of facial movement of each participant was plotted with respect to time, as well as the average across all participants (**Fig 6**). Individual reaction patterns appear distinct, suggesting that some participants may be more sensitive to certain images, and others less sensitive. The averages for each emotion demonstrate a consistent increase in facial movement throughout the viewing periods. Therefore, the general trend among participants reveals that the manifestation of happy and sad emotions in response to these images does not occur immediately, but rather builds and increases in magnitude until the viewing period concludes.

**Fig 6:**
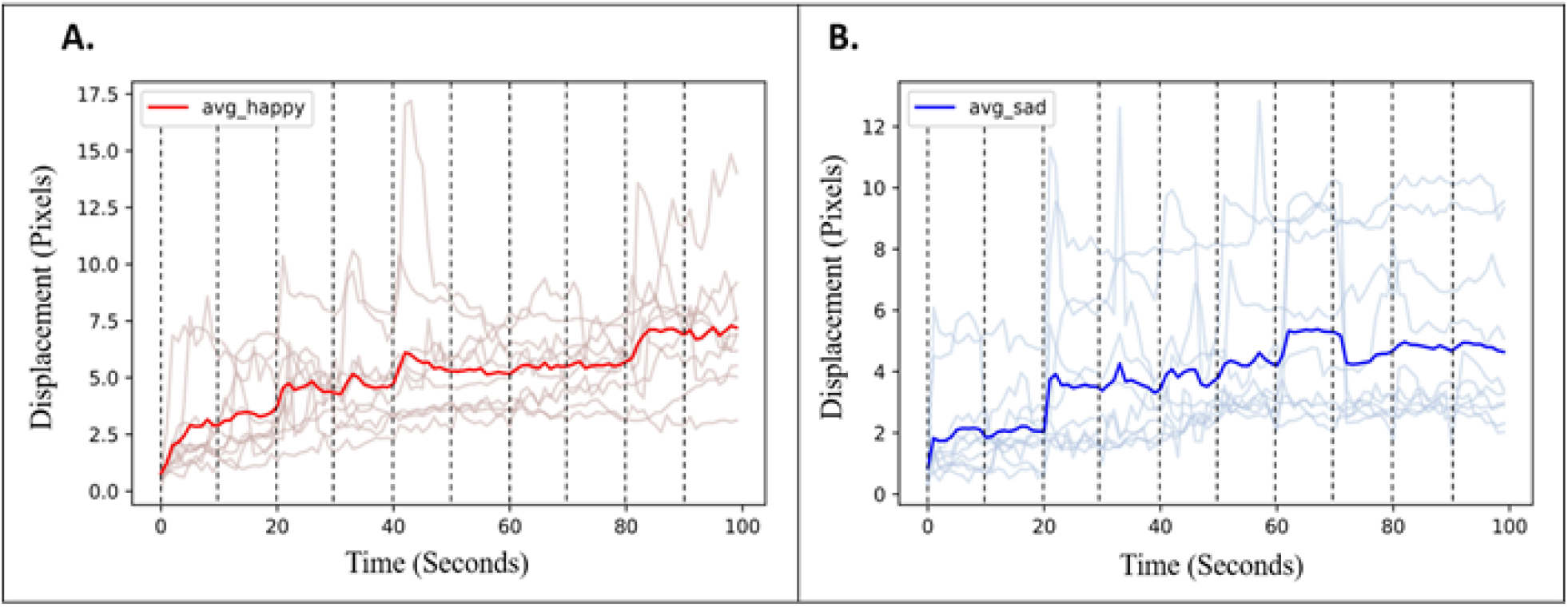
Temporal changes of average facial movement for each individual (ghosted lines) and across all participants (prominent line) over the duration of (A) happy and (B) sad image presentation. Dashed vertical lines represent the presentation of a new image.

### DISC Emotion Classifier

We constructed three machine learning classifiers that use information from the heatmaps generated by DISC to detect the emotional state of the individual. Each of the ten participants has 100 happy, 100 sad, and 30 neutral frames. The classifiers were trained on seven participants (7 × 230 = 1610 labeled images), and their performance was tested out-of-sample on the remaining three participants (3 × 230 = 690 labeled images). The three participants used out-of-sample had also provided informed consent for Amazon Rekognition analysis.

The out-of-sample classification results for each classifier are summarized in **Figure 7**. They consist of confusion matrices, where each row of the matrix represents the true emotion, and each column represents the predicted emotion. The numbers in the matrices are the percentage of the out-of-sample frames that an algorithm assigns to a given label. Each row sums up to 100%, and a perfect classification algorithm would have 100% in the rows and columns with the same labels. The trained classifiers performed a total of 690 (= 3 × 230) emotion predictions on the three test-set participants with 100 happy, 100 sad, and 30 neutral frames. All three DISC-based models were extraordinarily accurate, successfully predicting the participant’s emotion on 94-100% of frames (**Fig 7A-C**).

**Fig 7:**
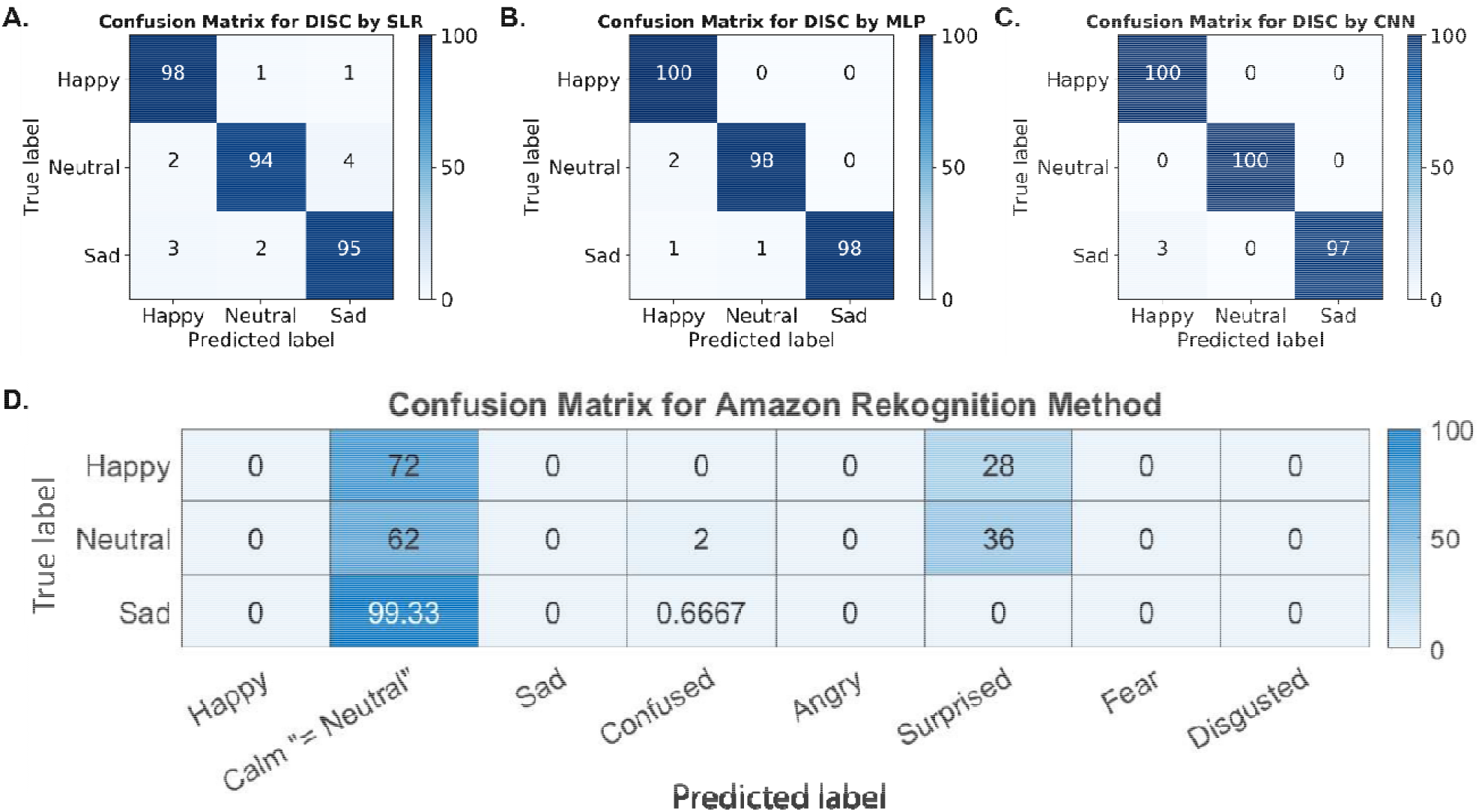
Confusion matrices of the two methods. (A)-(C): Different classifiers using the DISC method. (D): Amazon Rekognition. On the y-axis are the true states for the images; on the x-axis are the predicted states by the respective methods. The numbers in the plots indicate percentages of predicted states for each true state. Abbreviations: DISC (digital image speckle correlation), SLR (sparse logistic regression), MLP (multi-layer perceptron), CNN (convolutional neural network).

We also compared our three classifiers with the performance of Amazon Rekognition, a commercially available software that employs deep learning methods to analyze images of the face and detect emotions (**Fig 7D**). Interestingly, the Amazon Rekognition software classified a majority of the neutral, happy, and sad frames as calm or surprised. It was unable to both predict the correct mood and discriminate between happy, sad, or neutral frames, a distinction readily visible after DISC processing. Although the Amazon Rekognition method can classify images into eight different categories, it identified most of the frames as neutral. Hence, it is safe to conclude that DISC-based methods are more accurate in this case. Importantly, neither DISC nor Amazon Rekognition classifiers were trained on this data. We trained our DISC-based classifiers on frames from seven of our ten participants. Amazon Rekognition was pre-trained by AWS on a much larger database of images.

Nevertheless, our results show that by analyzing changes in facial expressions, DISC detects features that a static, state-of-the-art commercial emotion recognition tool cannot, and that these features are predictive of the participant’s underlying emotion.

## Discussion

In this study, we sought to establish proof-of-concept for DISC as a real-time emotional assessment tool. Research into the ability of various other biosignals [11–15,17–19] to measure emotion has gained popularity as these methods are dynamic and eliminate the need for self-report. The chief benefit of these biosignals is that they are responsive to stimuli, thus providing robust information. However, many of them are vulnerable to variability in measurement and interpretation, or lack a response sufficiently specific to reliably identify the underlying emotion. Analysis of facial movement via DISC mitigates these concerns by noninvasively monitoring changes in emotion in real-time. Furthermore, this technique differs from methods that track facial expressions with the FACS in that our participants did not display evidence of facial expression changes visible on raw video.

We presented healthy volunteers with a slideshow of images designed to trigger both happy and sad emotions while videotaping their faces. We utilized DISC to detect subtle facial movements during image viewing and we found that each image set reliably elicited spatially unique patterns of movement that were conserved across individuals. Moreover, the identified facial maps for happy- and sad-triggered emotions were invariant of participant age and gender. This observation is consistent with prior theories on the universal manifestation of emotions on the face [24,44]. In addition, PCA of our DISC movement data was capable of distinguishing happy and sad emotions in eight of our ten participants. This highlights the viability of our method in real-time emotion assessment simply by analyzing changes in facial expression undetectable to the naked eye.

When we investigated the temporal features of these subtle changes in facial expression, we discovered that the magnitude of movement increased throughout the viewing period. The idea that subtle facial movements can build throughout the experience of a particular emotional state distinguishes our observations from Ekman’s microexpressions, which are thought to last only a fraction of a second [27,45]. Whereas microexpressions were generally seen as a way to identify deception [28], the facial responses invisible to the naked eye observed in our study can be used to assess the participant’s emotion. We also observed that individual participants exhibited unique reaction patterns, with some showing spikes in facial movement at various points throughout the viewing period. This may be due to variations in an individual’s prior experiences and perceptions that influence the way they react to the images. Ultimately, further investigation into the temporal manifestation of emotion on the face is required to determine whether this observation holds true in a larger sample.

Finally, using the DISC-processed facial movement data, we built machine learning classifiers capable of predicting the emotion of individuals that they had not been trained on. The accuracy of our classifiers for any of the three emotion categories was between 94-100%. Comparison to the best commercially available emotion recognition software, Amazon Rekognition, revealed that DISC detects features that other deep learning methods simply cannot. Importantly, the features that our classifiers depend on for predictions are solely based on DISC facial movement output, whereas methods such as Amazon Rekognition use actual facial characteristics present in the images. This makes our method inherently unbiased with respect to analyzing individuals of different races or genders.

Other studies on tracking subtle changes in facial expression had participants falsely display or disguise their facial expressions or emotions [28,29,46]. In this study, we elicited organic reactions in the viewer through the presentation of images. These responses were then analyzed using DISC, followed by PCA and three different machine learning classifiers, thus maintaining the objectivity of our results. Aside from its objectivity, DISC analysis of emotion is exquisitely simple and cost effective, requiring only a digital camera and computer. Even the cameras on most smartphones can provide adequate resolution [32].

Among limitations to the current study is the potential for inducing the Hawthorne effect [47]. Participants were aware that they were being videotaped, which provided the opportunity for them to enhance their reactions to the images they viewed. Even so raw videos did not demonstrate overt changes in facial expression. Further testing including interspersing happy and sad images throughout the slideshow as well as introducing jittering to vary the length of image viewing are warranted to validate this method beyond our proof-of-concept investigation. Our small sample size notwithstanding, early results suggest promising applications to the fields of affective research and in clinical settings. With increased awareness of the need for patient-centered care, an objective tool for assessing emotions and mood would be immensely useful in monitoring the responses of patients with mood disorders to psychiatric treatment.

## Conclusion

Here we have demonstrated that individuals display subtle facial movements indicative of underlying emotions that are detectable with DISC. Our methodology has identified consistent facial maps for happy and sad emotions that are invariant of age and gender. PCA of the results of our facial movement data suggests that happy and sad emotions could be distinguished in as few as two dimensions. Our own machine learning algorithms can also use this data to reliably and accurately predict an individual’s underlying emotions and elucidate features of facial movement undetectable by a state-of-the-art emotion recognition software. Thus, our method demonstrates promise as an automated, noninvasive, quick-and-easy, affective assessment tool. We believe this tool can provide value in clinical monitoring settings as it has proven both robust and economical in predicting emotions.

## Data Availability

The data and code used for this manuscript are available upon request from the corresponding author. To protect participant privacy, raw videos of our participants cannot be shared publicly. Modified videos are available upon request from the corresponding author.

## Data and code availability

The data and code used for this manuscript are available upon request from the corresponding author. To protect our participants’ privacy, raw videos of our participants cannot be shared publicly. Modified videos are available upon request from the corresponding author.

## Credit author statement

**Jordan R. Saadon:** Conceptualization, Methodology, Investigation, Writing - Original Draft, Project Administration. **Fan Yang:** Software, Formal Analysis, Data Curation, Investigation, Writing - Original Draft, Visualization. **Ryan Burgert:** Software, Formal Analysis, Data Curation. **Selma Mohammad:** Software, Formal Analysis, Data Curation. **Theresa Gammel:** Investigation, Data Curation. **Michael Sepe:** Software, Formal Analysis. **Miriam Rafailovich:** Conceptualization, Resources, Writing - Review & Editing, Supervision. **Charles B. Mikell:** Conceptualization, Resources, Writing - Review & Editing, Supervision. **Pawel Polak:** Software, Formal Analysis, Writing - Review & Editing. **Sima Mofakham:** Conceptualization, Resources, Writing - Review & Editing, Supervision, Project Administration, Funding Acquisition.

## Acknowledgments

We would like to thank our volunteers for participating in this study as well as the Neurosurgery Department at Stony Brook University Hospital for their support.

## Supplementary Figures

**Supplementary Fig 1.**
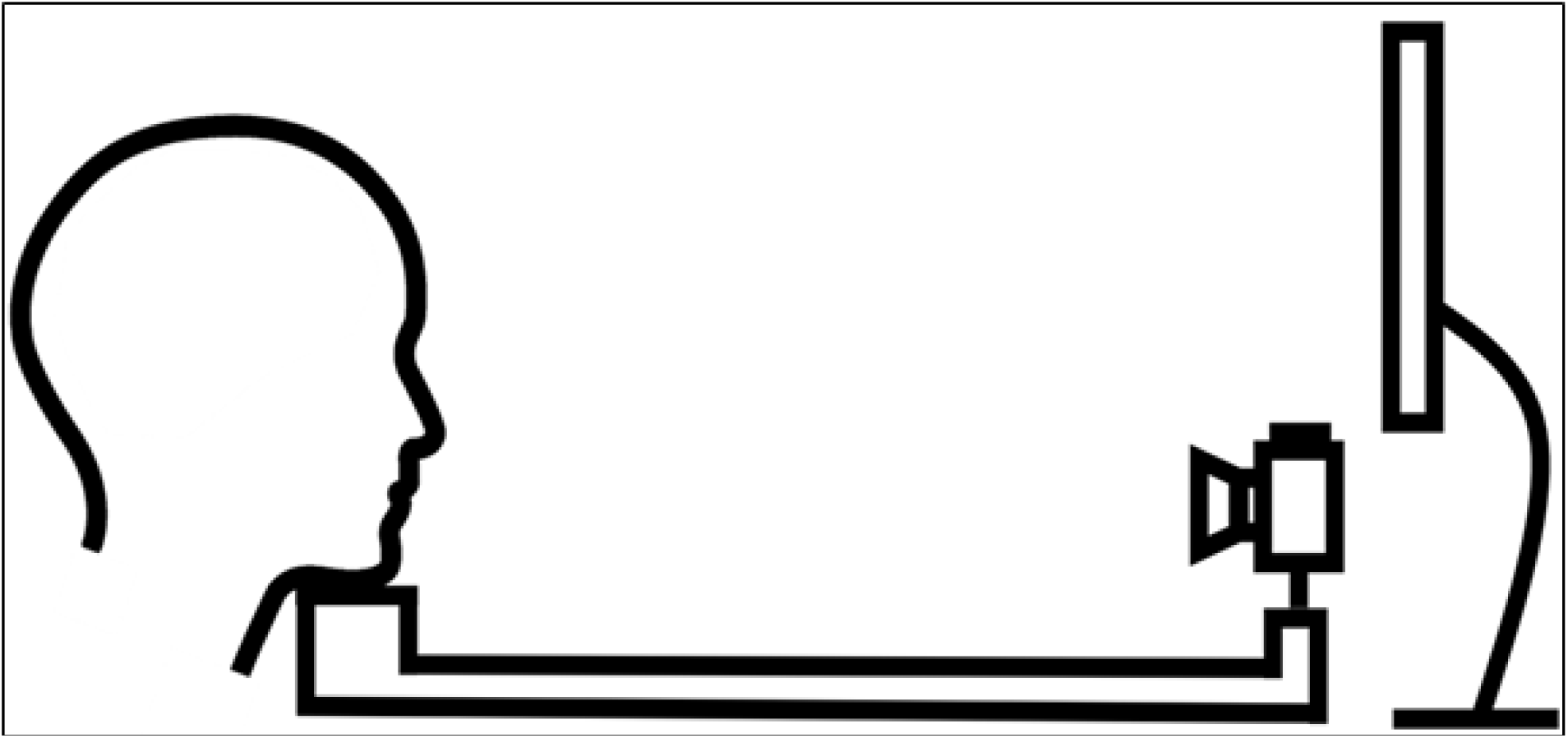
Schematic of the videotaping setup. Participants placed their chin on the chinrest, which is attached to the camera mount to allow recording of the face from a fixed distance. Behind the camera is the scree that presents the images during recording.

**Supplementary Fig 2.**
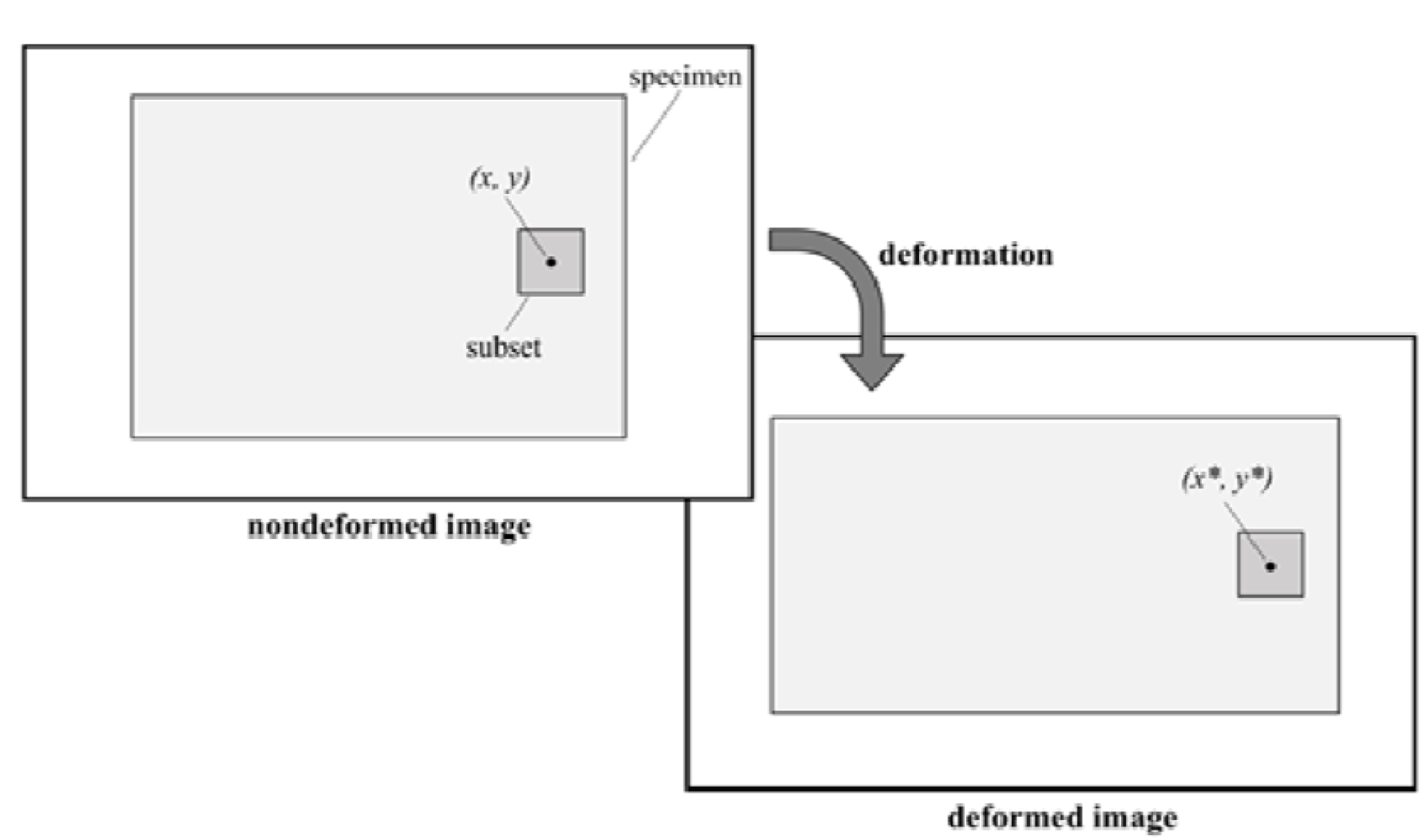
Schematic diagram of the DISC algorithm.

## Footnotes

1 We modified the following publicly available code for our analysis: https://gitlab.com/damien.andre/pydic

2 Software is available at: https://aws.amazon.com/rekognition

3 This figure was removed from the manuscript because it contains images of our participants’ faces, per medRxiv policy. Please contact the corresponding author for access to these images.

4 This figure was removed from the manuscript because it contains images of our participants’ faces, per medRxiv policy. Please contact the corresponding author for access to these images.

